# Longitudinal enlargement of choroid plexus is associated with inflammation and neurodegeneration in RRMS patients

**DOI:** 10.1101/2023.08.06.23293738

**Authors:** Samuel Klistorner, Alexander Klistorner

**Author notes:** Corresponding author: Alexander Klistorner.

## Abstract

**Background:** The Choroid Plexus (CP) plays a potential role in the initiation and propagation of neuroinflammatory processes in MS. However, the progressive change of the CP and its associations with biomarkers of acute and chronic inflammation, along with MS-related brain structure atrophy, have not been investigated.

**Objective:** This study aims to explore the longitudinal dynamics of the CP in RRMS patients and assess its relationship with inflammation and with atrophy in various brain compartments.

**Methods:** 57 RRMS patients were examined annually for a minimum of 60 months using following MRI protocols: pre- and post-contrast (gadolinium) Sagittal 3D T1, FLAIR CUBE, diffusion weighted MRI. CP was manually segmented at baseline and last follow-up and normalised by head size. Annually lesion segmentation was performed by iQ-MSTM software suite and brain was segmented using AssemblyNet.

**Results:** Over the study period, the volume of CP increased by an average of 1.4% annually. The magnitude of CP enlargement significantly correlated with central brain atrophy, and to a lesser extent, total brain atrophy, white matter, and deep grey matter atrophy. Furthermore, progressive CP enlargement was significantly associated with the volume and severity of chronic lesion expansion (r=0.66, p<0.001), but not with the number or volume of new lesions.

**Conclusion:** Our findings indicate that ongoing inflammatory activity in the CP is linked to low-grade demyelination at the rim of chronic lesions and associated neurodegeneration of periventricular white and grey matter.

## Introduction

Multiple sclerosis (MS) is a chronic neurodegenerative disease characterized by the presence of inflammatory demyelinating lesions in the central nervous system (CNS). The progression of MS is complex and involves a combination of immune-mediated inflammation and neurodegenerative processes. Acute MS lesion formation is characterized by disruption of the blood-brain barrier (BBB) and infiltration of adaptive immune cells and monocytes.

However, concomitant with this acute process, a gradual accumulation of low-grade inflammation occurs within the central nervous system (CNS), referred to as compartmentalized inflammation. This “slow-burning” inflammatory demyelination at the periphery of chronic MS lesions plays a crucial role in lesion expansion over time and is implicated in disease progression, including neurodegeneration, brain atrophy, and worsening disability.[1][2][3][4]

Recent studies have also shed light on the role of the choroid plexus (CP), a highly vascularized structure within the brain ventricles, in the pathophysiology of MS. [5][6][7][8][9][10][11] The choroid plexus plays a critical role in regulating the composition of cerebrospinal fluid (CSF) and facilitating the entry of immune cells into the CNS. It has also been implicated in the initiation and propagation of neuroinflammatory processes in MS.

A recent cross-sectional study involving MS patients revealed an intriguing correlation between the size of the CP and the degree of chronic lesion expansion. [8] These findings suggest that alterations in the CP may be linked to the progression of chronic lesions in MS. In contrast, a separate study focusing on patients with clinically isolated syndrome (CIS), an early stage of MS, demonstrated a transient increase in the size of the CP during new bouts of acute inflammation.[9] This suggests a dynamic relationship between the CP and acute inflammatory processes in the CNS.

Given these findings, there is a need to investigate the longitudinal changes in the CP among relapsing-remitting MS (RRMS) patients and explore its potential association with both acute and chronic inflammation and neurodegeneration. Therefore, the aim of the current study is to examine dynamics of the CP in RRMS patients and assess its relationship with markers of inflammation and neurodegeneration.

## Methods

The study was approved by the University of Sydney and Macquarie University Human Research Ethics Committees and followed the tenets of the Declaration of Helsinki. Written informed consent was obtained from all participants.

### Subjects

Patients diagnosed with RRMS according to the revised McDonald 2017 criteria[12] who were enrolled in an on-going longitudinal study of MS-related axonal loss and who completed at least 4 years follow-up were included in the study. Patients underwent annual MRI scans and clinical assessment.

### MRI protocol and analysis

MRI was performed using a 3T GE Discovery MR750 scanner (GE Medical Systems, Milwaukee, WI). The following MRI sequences were acquired: Pre- and post-contrast (gadolinium) Sagittal 3D T1, FLAIR CUBE, diffusion weighted MRI.

Specific acquisition parameters and MRI image processing are described in more detail in Supplementary Material

Diagram demonstrating the flow of MRI processing and the specific type of analysis they were used for is shown in Fig.1

**Fig. 1.**
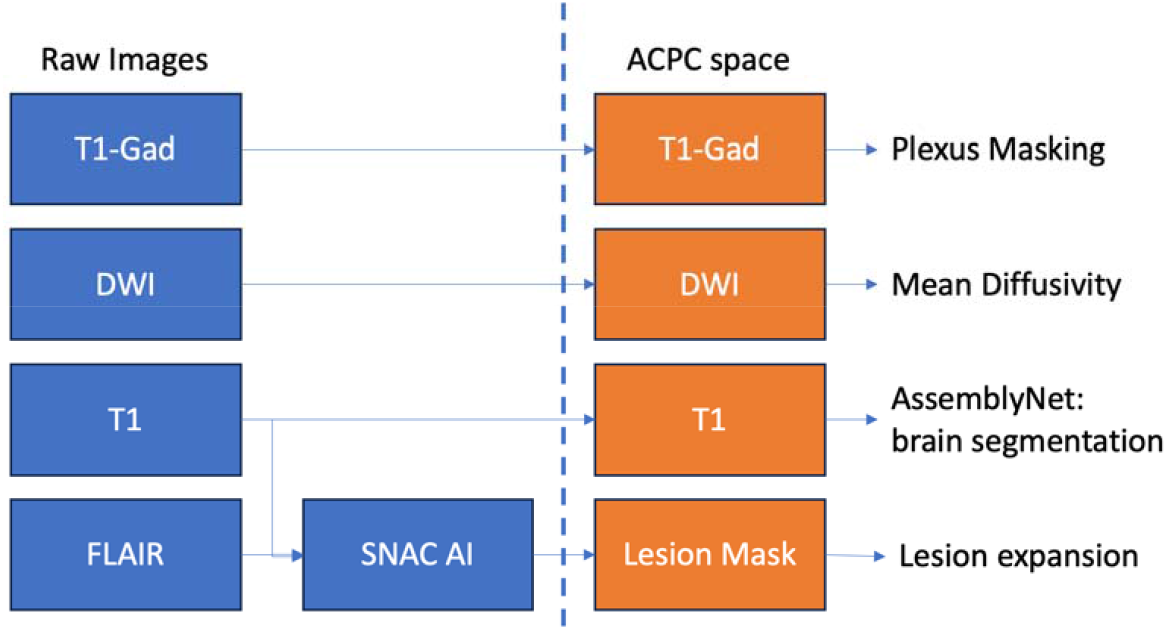
Diagram demonstrating the flow of MRI processing. Note that lesion segmentation was performed using T1 and FLAIR images in native space, while all other analysis-in ACPC space.

#### Longitudinal CP analysis

The CP within lateral ventricles was manually segmented on ACPC co-registered T1 Gad-enhanced images (which represents the gold-standard technique to non-invasively segment CP [13]) (Fig.2, upper row) using JIM 9 software (Xinapse Systems, Essex, UK) by a trained analyst (AK) at the baseline and last follow-up visits. The analyst was blinded to both clinical and lesion data.

**Fig. 2.**
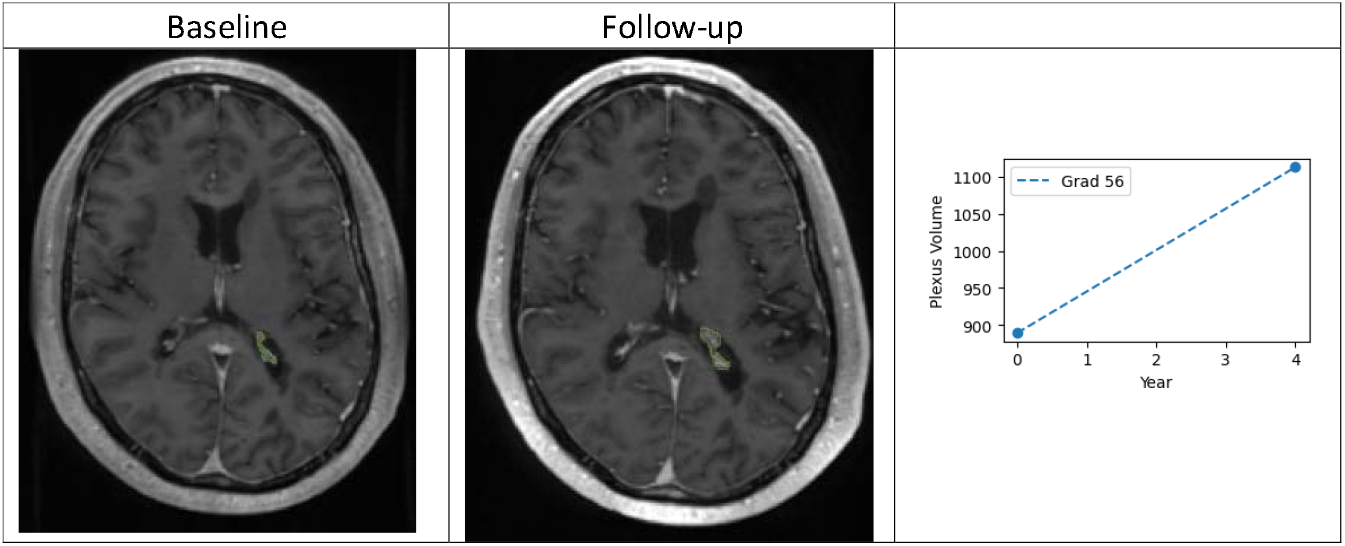
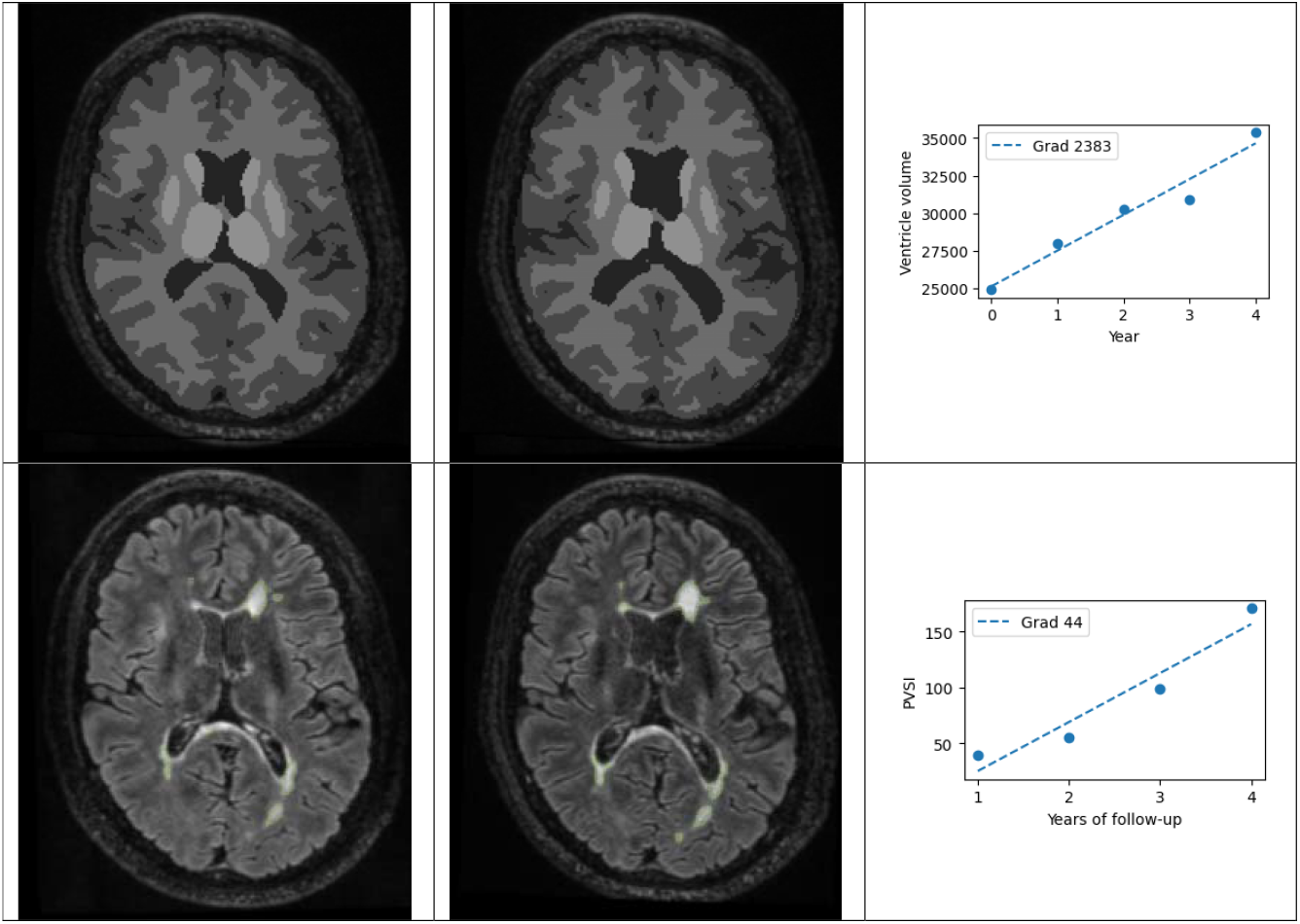
Example of plexus delineation using manual segmentation (only right side shown for clarity) (upper row), brain segmentation using AssemblyNet (middle row) and lesional segmentation using iQ-MSTM (bottom row). All mages presented at baseline and last follow-up (48 months). Right column demonstrates longitudinal values of plexus volume (upper row), ventricle volume (middle row) and chronic lesions volume (bottom row) annual. Best fitting line represents annual gradient of change.

To account for inter-subject variability of head size, the CP volume was normalised (CPn) using SienaX-based scaling, as described previously. [9] The average annual normalised CPn volume change was calculated for each patient.

#### Lesion analysis

T2 lesions were segmented for all time points using a fully automated lesion segmentation algorithm from the iQ-MSTM software suite (Sydney Neuroimaging Analysis Centre, Sydney, Australia), using unprocessed T1 and FLAIR images. These lesion masks were then transformed to ACPC space using linear registration. Only lesions >50mm3 were included in analysis.

For each subject, lesion masks for each pair of neighbouring time-point were then processed through our in-house software to determine the degree of chronic lesion expansion between consecutive time-points while correcting for brain atrophy-related displacement of lesions, as described previously (Fig.2 bottom row).[4] Cumulative volume of lesion expansion for each patient was then computed by summarizing annual expansion values. To account for difference in duration of follow-up between subjects an average annual lesion expansion volume was used for analysis (Fig.2 bottom row).

The degree of tissue damage inside the lesions and within the lesion’s expanding part was measured as an increase of Mean Diffusivity (MD) between baseline and follow-up timepoints, as proposed before.[14] To adjust for the severity of damage inside the expanding part of chronic lesions a progressive volume/severity index (PVSI) was calculated by multiplying volume of lesion expansion by change of MD in the corresponding (expanding) area, as described earlier.[15] Progressive tissue destruction inside chronic lesions was measured as an average annual increase of MD using the baseline lesion mask, which was adjusted to correct for brain atrophy-related displacement of lesions at follow-up, as described previously.[14]

#### Identification of new acute lesions

This study focused on the analysis of the relationship between CP enlargement and chronic and acute lesions. Therefore, it was necessary to distinguish between the two types of lesions. As a result, additional steps were taken to identify and differentiate acute lesions that occurred during the study period.

All gadolinium-enhancing lesions occurring at any time point were considered “new lesions” and were excluded from the analysis of chronic lesion expansion for the next annual interval. However, gadolinium enhancement within active MS lesions usually does not persist beyond 2 months, after which newly formed T2 hyperintense lesions continue to shrink in size for another 3-5 months, reflecting resolution of edema and, potentially, tissue repair including remyelination [16]. Therefore, to accurately identify new lesions, we examined all lesions shrinking more than 20% between two consecutive time-points. We determined whether these lesions were newly formed at the previous time-point or already existing. All newly formed lesions were classified as ‘acute lesions’ and were only included in the analysis of chronic lesions from the next time-point onwards.

##### Volumetric brain analysis

Volumetric measures were obtained using AssemblyNet, an AI brain segmentation tool,(Coupé et al., 2020) on T1 images in ACPC space.

Following metrics was analysed: total brain atrophy, white matter atrophy, grey matter atrophy, cortex and deep grey matter atrophy and ventricular volume change. For this study, volumetric change of ventricles was used as a measure of central brain atrophy (CBA).^16 17^ (Fig.2 middle row)

The cerebellar was excluded from the white and grey matter analysis.

To account for variability in the number of follow-up years between subjects, we established “a volume change per year” metric as a measure of atrophy. This involved calculating the annual change for each MRI measure and generating a line of best fit to represent the annual changes. The gradient of the line of best fit represented the annual change (see for instance Fig. 2 middle row, right panel).

### Statistics

Statistical analysis was performed using SPSS 22.0 (SPSS, Chicago, IL, USA). Pearson correlation coefficient was used to measure statistical dependence between two numerical variables. For partial correlation, data was adjusted for age, gender and disease duration. Average annual volume of new lesions was added to partial correlation for analysis of relationship between CP change and expansion of chronic lesions. P < 0.05 was considered statistically significant. Shapiro–Wilk test was used to test for normal distribution.

To assess the significance of longitudinal changes while accounting for variation in the duration of follow-up between patients, a linear mixed-effects model was employed. In this model, the volume measure of interest (CPn, lesions or various metrics of brain atrophy), collected at two time points (baseline and follow-up) serve as a dependent variable. The fixed effects of the model comprised time (baseline vs. follow-up), while controlling for age, sex and disease duration. The random effects included the individual subjects to account for the repeated measures on the same subjects over time.

## Result

There were 57 patients who satisfied inclusion criteria. Demographic data is presented in Table 1.

| Demographic Variable |  |
| --- | --- |
| Total number of patients | 57 |
| Age at enrolment, years | 40.4±9.0 |
| Gender | 22M/35F |
| Disease duration at enrolment, years | 6.9±5.9 |
| Duration of the follow-up, months | 69.1+/-15.4 months |
| EDSS (median) | 1.1 |

In our RRMS cohort, the baseline volume of CPn was measured at 1308+/-423 mm^3^ (Fig.3 a). Over the study period, the volume of CPn increased significantly (p<0.00001, mixed-effect model). The average annual CPn volume enlargement was 19.1+/-17.8 mm^3^ or 1.4+/-1.2% (Fig.3 b, c) However, the extent of enlargement varied considerably among individuals, ranging from -1.8 mm3 to 75.4 mm^3^ or -0.2% to 6.3% (Fig. 3). Example of longitudinal CP segmentation is presented in Fig.2 (upper row)

**Fig. 3.**
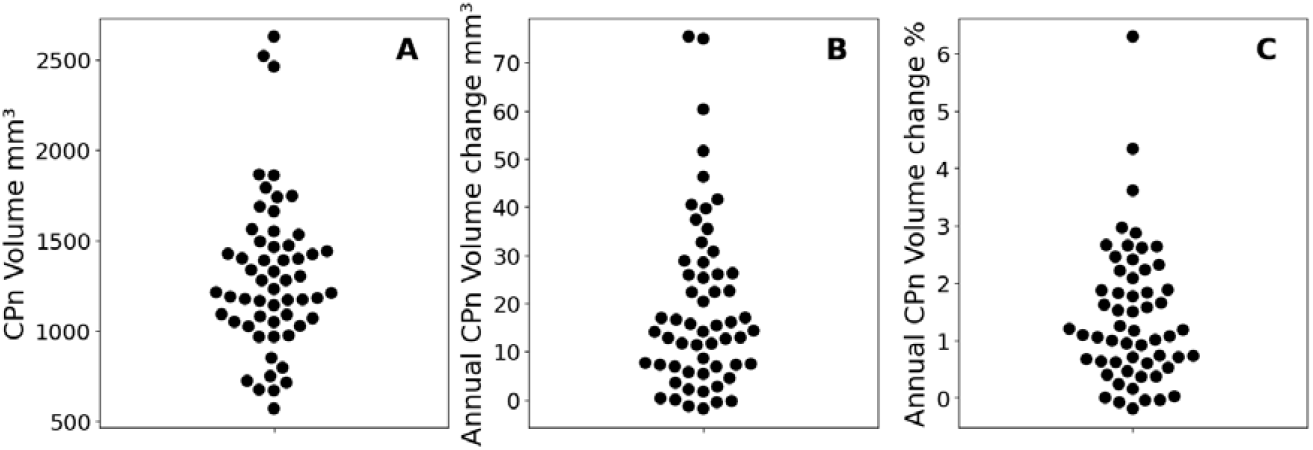
Distribution of CPn volume at baseline (a), absolute (b) and relative (c) annual increase of CPn volume.

Volume of CPn enlargement during follow-up correlated significantly with the size of the plexus at baseline (r=0.50, p<0.001) (Fig 4a), but showed no association with gender, age, disease duration or EDSS.

**Fig. 4.**
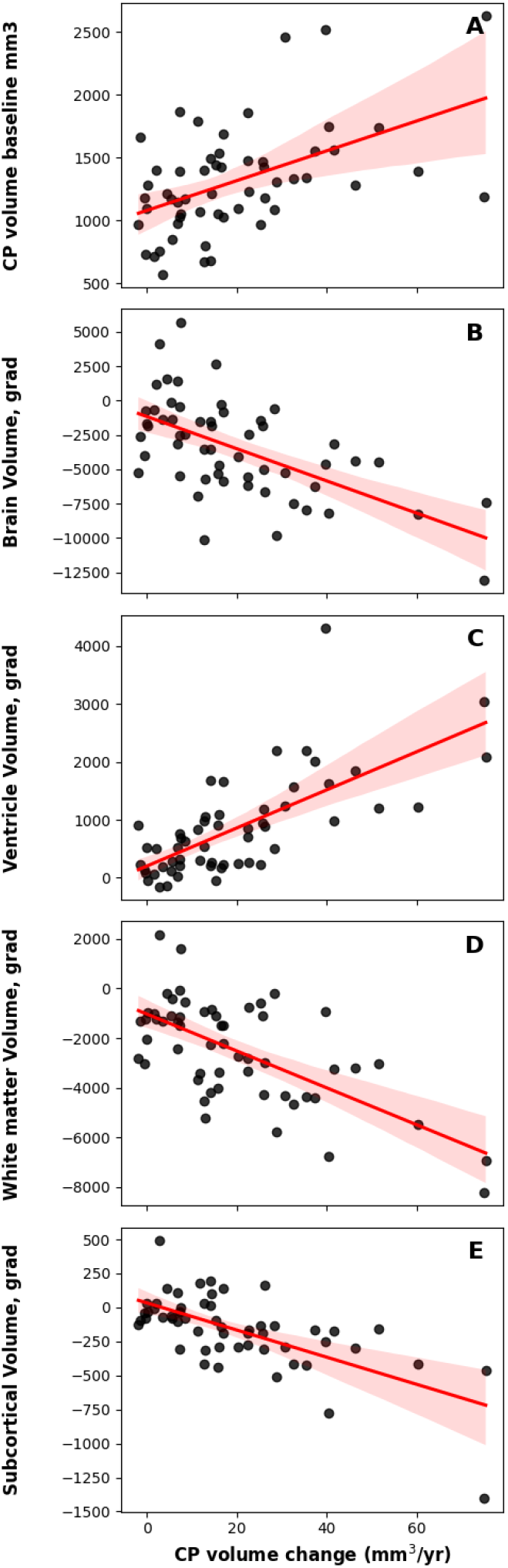
Correlations between CPn volume change and longitudinal brain volumetrics. Shaded area represents confidence interval.

We found statistically significant total brain atrophy (0.23% per year), including white matter (0.48% per year), total grey matter (0.12% per year), deep grey matter (0.29% per year) and cortex (0.11% per year). The observed atrophy in central brain (ventricular volume), however, was notably larger (2.68% per year) (p<0.001 for all, mixed-effect model). Disease duration had significant effect on total brain atrophy (p=0.001), ventricular volume (p=0.006), white and deep grey matter atrophy (p<0.001 and 0.01 respectively), while age contributed to total brain atrophy (p=0.037), grey matter atrophy (p=0.002) and cortex (p=0.001).

We also found substantial increase of total T2 lesion load during the study period (4621±4952 mm^3^ and 6076±5981 mm^3^ at baseline and last follow-up respectively, p<0.0001, mixed-effect model). (Duration of follow-up contributed significantly to the model, p<0.001).

There were on average 0.3±0.5 new lesions identified annually (average annual volume of new lesions 45.2±90.0 mm^3^).

Furthermore, we observed substantial expansion of chronic lesions (346±449mm^3^ average per year), which demonstrated considerable inter-subject variability (coefficient of variability 130%).

Baseline value and volumetric change per year are presented in Table. 2

**Table 2.** Brain volumetric data and annual change.

|  | Total brain volume | White matter volume | Grey matter volume | Cortical volume | Deep grey matter volume | Ventricle volume | CPn |
| --- | --- | --- | --- | --- | --- | --- | --- |
| Baseline | 1577697<br>+/-51330 | 533337<br>+/-33353 | 842075<br>+/-25373 | 784786<br>+/-24026 | 57289<br>+/-3854 | 31640<br>+/-14435 | 1308 +/-423 |
| Change per year | -3599<br>+/-3741 | 2522<br>+/-2066 | 1046<br>+/-1978 | 883<br>+/-1860 | 163<br>+/-270 | 877<br>+/-880 | 19.1 +/-17.8 |

### Association of CPn change with longitudinal brain volumetric parameters

A significant correlation was observed between the annual CPn volume increase and the annual rate of total brain atrophy (r=-0.59, p<0.001, partial correlation: r=-0.59, p<0.001) (Fig 4b). This association noticeably strengthened when central brain atrophy (as measured by ventricle enlargement) was considered in isolation (r=0.70, p<0.001, partial correlation: r=0.71, p<0.001) (Fig 4c).

CPn enlargement also exhibited a moderate correlation with the annual rate of white matter atrophy (r=-0.65, p<0.001, partial correlation: r=-0.64, p<0.001) (Fig 4d). The link between CPn enlargement and grey matter atrophy, however, was weak (r=-0.34, p=0.012, partial correlation: r=-0.38, p=0.008) and was mainly driven by deep grey matter (r=0.66, p<0.001, partial correlation: r=-0.65, p<0.001) (Fig 4e), since the association with cortical grey matter demonstrated borderline significance (r=-0.25, p=0.06, partial correlation: r=-0.28, p=0.04).

### Association with lesions

In our study, the annual volume of CP enlargement significantly correlated with the volume of chronic lesion expansion (r=0.47, p<0.001, partial correlation adjusted for age, sex, disease duration and annual volume of new lesions) (Fig 5a). The strength of this association markedly increased when the degree of tissue damage within expanding part of the lesion, in the form of the progressive volume/severity index (PVSI)[15], was considered (r=0.66, p<0.001, partial correlation: r=0.64, p<0.001) (Fig. 5b). CP enlargement was also linked to the degree of tissue rarefication inside chronic lesions, as measured by MD increase (r=0.50, p<0.001).

**Fig. 5.**
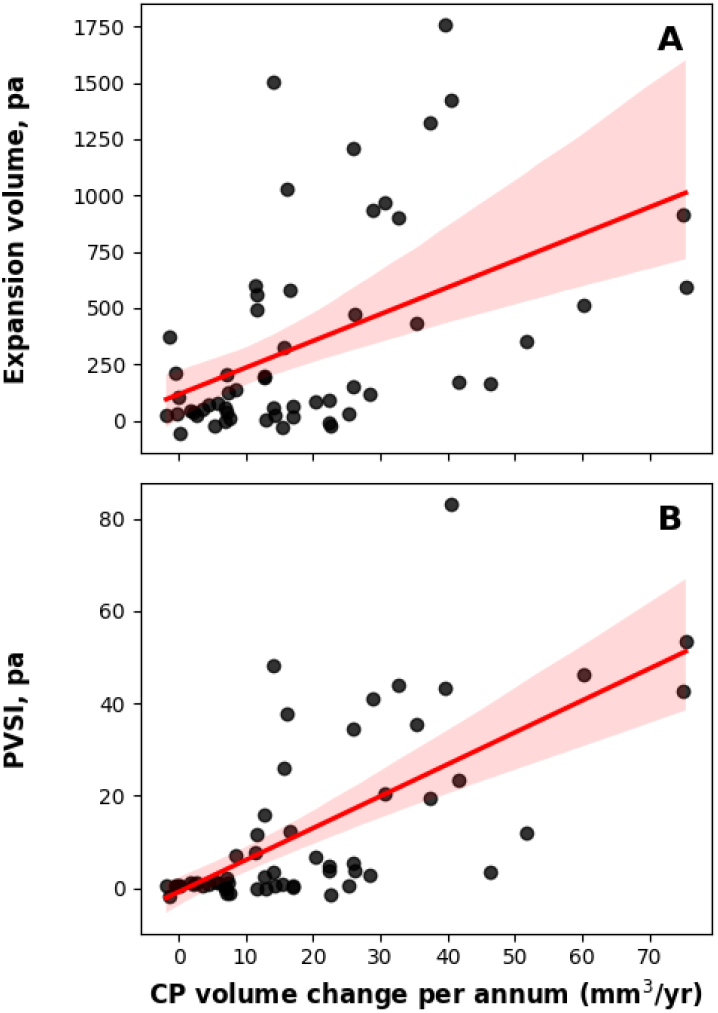
Correlation between CPn volume change and volume (a) and PVSI (b) of chronic lesion expansion.

However, no significant relationship was found between changes in CP and the number or volume of new lesions.

## Discussion

This study aimed to investigate progressive change of CP in RRMS patients and its associations with biomarkers of both acute and chronic inflammation as well as MS-related brain atrophy.

For the first time, our study has established that the volume of CP in RRMS patients does, in fact, increase gradually over the course of the disease. In our patient cohort, we found that the average annual rate of CP enlargement was 1.4%, though there was a wide individual range of change from -0.2% to 6.3% (equivalent to -1.8 mm^3^ to 75.4 mm^3^).

There are several aspects of the study’s design that enabled us to detect such a subtle change. The utilization of manual segmentation, despite being a laborious and very time-consuming process, was critical in achieving high-quality delineation of the CP. Additionally, the extended duration of follow-up period, lasting up to 8 years, significantly contributed to the robustness of our findings. The lengthy observation time enabled more accurate estimation of CP change by effectively minimizing the noise of measurement. As a result, it contrasts with our previous study[18], in which we were unable to detect changes in CP during a 4-year observation period, thereby indicating the importance of long-term observation in capturing slow and gradual CP volume changes in RRMS.

### Increase of CP volume and inflammation

The vital role of the choroid plexus in maintaining the blood–CSF barrier and modulation of inflammatory cells trafficking into the central nervous system has recently attracted the attention of MS researchers.[19][20] It has become clear that the plexus not only actively participates in acute inflammatory processes including antigen presentation and recruitment of peripheral inflammatory cells,[21] but remains chronically inflamed, even in long-standing MS,[22] contributing, therefore, to a proinflammatory state of the CSF and inducing a persistent neuroinflammatory environment in periventricular brain tissue.[23].

Several recent cross-sectional studies have demonstrated significantly larger CP in patients with MS compared to normal controls, even at early stages of the disease.[24][7][18][19][9][10] Various hypotheses, such as CSF hypersecretion,[25] oxidative stress,[26] or edema[27] have been proposed to explain CP enlargement in MS. However, numerous experimental and imaging studies have demonstrated a close association between parenchymal inflammation of plexus tissue and increases in CP volume, suggesting inflammation as the likely cause of this enlargement. [22][28][5][7][13][24] Therefore, we speculated that the broad range of CP enlargement observed in our study likely reflects varying degrees of plexus inflammation in our RRMS cohort.

This notion aligns with another significant finding of this study - the observed link between progressive CP enlargement and the volume and severity of chronic lesion expansion. This association suggests that the CP could be indirectly implicated in forming/sustaining the slow-burning inflammatory process at the rim of chronic periventricular MS lesions, possibly through the secretion of pro-inflammatory cytokines or the regulation of immune cell trafficking and microglial activation[29][30] [31] This view is supported by our earlier study which demonstrated a clear link between the size of the CP and the rate of chronic lesion expansion in patients with RRMS[18] and also aligns with recent publication demonstrating larger CP volume in patients with chronic active lesions. [28]

Moreover, it may partially explain the predominantly periventricular effect of CP enlargement on brain atrophy. While our results show that CP enlargement significantly correlate with atrophy of both white and grey matter, supporting the view that inflammation originating in the CP may contribute to ongoing neurodegeneration, these associations are considerably stronger in the central brain (ventricle enlargement *vs* total brain atrophy or deep grey matter loss *vs* cortex atrophy). This suggests that these relationships are largely driven by the periventricular region, which also demonstrates markedly faster rate of both white and grey matter atrophy compared to brain periphery.

Interestingly, our recent study demonstrated that the extent of chronic lesion expansion (an imaging equivalent of low-grade inflammation at the lesion’s edge) also exhibits a clear CSF-related gradient, being more pronounced in the periventricular region. It is understood that this slow-burning inflammatory demyelination results in transection of some of the axons traversing the chronically inflamed lesion rim, thus leading to retrograde and anterograde (Wallerian) axonal degeneration. This has potentially detrimental effects on anatomically interconnected areas of the brain, leading to subsequent brain atrophy.[32] As stated earlier, since this process predominantly occurs in proximity to the CSF, it is not surprising that the periventricular area of the brain (and specifically the periventricular white matter and deep grey matter) is the most impacted. Therefore, any pathological influence exerted by the CP on the extent and severity of tissue damage at the rim of chronic periventricular lesions will directly contribute to the loss of periventricular axons and neurons and, consequently, to the shrinkage of periventricular white and grey matter. This is also in line with our previous findings linking CBA almost exclusively with the expansion of chronic lesions.[15]

Furthermore, the significant association of CP enlargement with progressive tissue rarefication inside chronic lesions, as measured by the increase of MD, also strengthens the link between change in plexus volume and tissue damage related to axonal transection at the lesion rim [4],[33].

A periventricular gradient of tissue damage has recently been described in normal-appearing and lesional tissue of both white and grey matter[34] for review). [35]. [36] [37]. It was suggested that this CSF-related gradient of damage is driven by a mechanism linked to microglial activation and/or an increased release of proinflammatory and cytotoxic cytokines from immune cell aggregates into the CSF. [34] [30] It has been proposed that CSF inflammatory/cytotoxic factors may induce diffuse pathological cell alterations directly, by mediating neuroaxonal injury, and/or indirectly, by activating tissue resident microglia and astrocytes.[36] Similar periventricular gradient of microglial activation and microstructural tissue damage has also been recently shown in MS patients in vivo. Compared to the white matter of heathy controls, patients with MS show a higher percentage of DPA+ voxels (indicating activated innate immune cells) at the closest proximity to the ventricles, which was associated with a corresponding increase of MTR. [29]

Periventricular gradient of neuroaxonal damage described in deep grey matter was also shown to be inversely associated with microglial activation and influenced by the presence of intrathecal inflammation.[36] Given the close proximity of subcortical nuclei to the ventricles (and therefore, to the CP), the observed deep grey matter damage might originate from direct influence of plexus-induced inflammation or, alternatively, from axonal transection taking place within the rim of chronic expanding lesions, which ultimately results in neuronal death. However, the second option appears more plausible since, contrary to the deep grey matter, we found no association between CP enlargement and cortical volume. Our finding of a differential impact of CP enlargement on deep and cortical grey matter is corroborated by recent research from Wang et al.[[28] who also established an association between enlarged CP and reduced volumes of deep grey matter, but found no such relationship with cortical volume.

Notably, we did not find a significant relationship between CP enlargement and the number or volume of new lesions. This observation could suggest that the role of CP might be more related to the chronic, rather than the acute phase of the disease. This, however, warrants further investigation since earlier study demonstrated weak, but significant association between the size of CP and new lesional activity [24].

Taken together, our study unequivocally establishes a link between the longitudinal enlargement of CP volume and various measures of neurodegeneration, including axonal loss in the rim and body of chronic lesions, as well as periventricular brain atrophy. Remarkably, while axonal damage related to chronic lesions and the atrophy of the brain’s white and grey matter were derived using different techniques of image analysis, they showed similar associations with CP enlargement, increasing robustness of our findings.

Building on existing literature on this subject, we propose the following model as a possible interpretation of our findings: increased inflammation of the CP triggers periventricular microglial activation and/or prompts the release of pro-inflammatory and toxic cytokines from the CSF into periventricular white matter (Fig.6, point 1), This, subsequently, activates/sustains a low-grade inflammation at the edges of chronic periventricular lesions, leading to axonal transection and lesion expansion (Fig.6, point 2). The ensuing retrograde and anterograde (Wallerian) degeneration of axons transected in the expanding areas of the affected lesions (Fig.6, point 3) ultimately results in predominantly periventricular white matter atrophy (Fig.6, point 4) and deep grey matter loss (Fig.6, point 5). However, while this model aligns with our data, it remains largely conjectural and requires verification through experimental research.

**Fig. 6.**
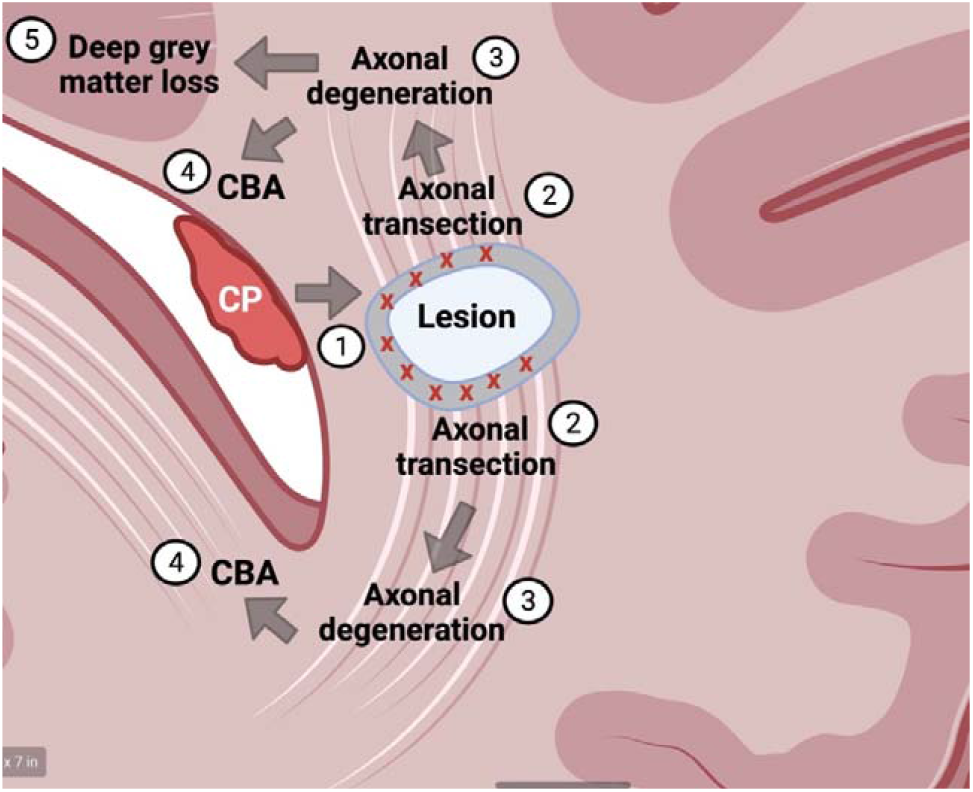
Diagram showing hypothetical sequence of event triggered by CP inflammation. 1. increased inflammation of the CP triggers periventricular microglial activation and/or prompts the release of pro-inflammatory and toxic cytokines from the CSF into periventricular white matter 2. axonal transection (red crosses) and lesion expansion (grey area around the lesion) 3. retrograde and anterograde (Wallerian) degeneration of axons transected in the expanding areas of the affected lesions 4. periventricular white matter atrophy 5. deep grey matter loss

There are several limitations to this study. First, our analysis was based on a relatively small sample of RRMS patients. Larger studies are necessary to confirm these findings and further elucidate the nature of the observed associations.

Secondly, our study was not designed to directly measure inflammation within the CP or brain tissue, and we used volume changes and lesion dynamics as indirect indicators of these processes. More direct measures of inflammation, such as the use of positron emission tomography (PET) with tracers targeting activated microglia or other inflammation-specific markers,[24] may provide a more comprehensive understanding of the relationship between CP changes and inflammation.

Thirdly, our study, while longitudinal, was observational in nature. As a result, while we can draw associations from the data, we cannot make causal inferences. It is not clear from our study whether CP enlargement directly contributes to inflammation at the rim of chronic lesions and subsequent brain atrophy, or whether they simply occur concurrently due to a shared underlying mechanism.

Lastly, while we used manual segmentation for a more accurate estimation of CP volume, this process has its own limitations, including being labor-intensive and subject to human error. Furthermore, it’s not feasible for routine clinical use or large-scale studies. The development of automated or semi-automated segmentation methods for CP could potentially overcome this limitation, enabling more widespread and efficient analysis of CP changes in RRMS patients.

In conclusion, this study provides the first evidence of progressive CP enlargement in RRMS patients and its association with chronic lesion expansion and brain atrophy, supporting a potential role for the CP in the chronic inflammatory processes and neurodegeneration in RRMS.

## Data Availability

All data produced in the present study are available upon reasonable request to the authors

## References

1. Calvi A, Carrasco FP, Tur C, et al. Association of Slowly Expanding Lesions on MRI With Disability in People With Secondary Progressive Multiple Sclerosis. 2022.

2. Dal-Bianco A, Grabner G, Kronnerwetter C, et al. Slow expansion of multiple sclerosis iron rim lesionsllJ: pathology and 7 T magnetic resonance imaging. Acta Neuropathol. 2017; 133(1):25–42.

3. Elliott C, Belachew S, Wolinsky JS, et al. Chronic white matter lesion activity predicts clinical progression in primary progressive multiple sclerosis. Brain. 2019; 142(9):2787–2799.

4. Klistorner S, Barnett MH, Yiannikas C, et al. Expansion of chronic lesions is linked to disease progression in relapsing–remitting multiple sclerosis patients. Multiple Sclerosis Journal. 2021; 27(10):1533–1542.

5. Fleischer V, Gonzalez-Escamilla G, Ciolac D, et al. Translational value of choroid plexus imaging for tracking neuroinflammation in mice and humans. Proc Natl Acad Sci U S A. 2021; 118(36):1–12.

6. Ricigliano VA, Morena E, Colombi A, et al. Choroid Plexus Enlargement in Inflammatory Multiple sclerosis. Radiology. 2021; 301:166–177.

7. Manouchehri N, Stüve O. Choroid plexus volumetrics and brain inflammation in multiple sclerosis. Proc Natl Acad Sci U S A. 2021; 118(40):10–12.

8. Klistorner S, Barnett M, Parratt J, et al. Choroid Plexus Volume Predicts Expansion of Chronic Lesions and Associated Brain Atrophy in Multiple Sclerosis. Ann Clin Transl Neurol. 2022; ccepted.

9. Klistorner S, Van der Walt A, Barnett MH, et al. Choroid plexus volume is enlarged in clinically isolated syndrome patients with optic neuritis. MSJ. 2022:Online ahead of print.

10. Ricigliano VAG, Louapre C, Poirion E, et al. Imaging Characteristics of Choroid Plexuses in Presymptomatic Multiple Sclerosis: A Retrospective Study. Neurol Neuroimmunol Neuroinflamm. 2022; 9(6):1–10.

11. Muller J, Sinnecker T, Wendebourg M, Schlger M, Yaldizi O. Choroid Plexus Volume in Multiple Sclerosis vs Neuromyelitis Optica Spectrum Disorder. Neurol Neuroimmunol Neuroinflam. 2022; 9:e1147.

12. Thompson a. J, Banwell B, Barkhof F, et al. Diagnosis of multiple sclerosis: 2017 revisions of the McDonald criteria. The Lancet (Neurol). 2018; 17:162–173.

13. Muthuraman M, Oshaghi M, Fleischer V, et al. Choroid plexus imaging to track neuroinflammation-a translational model for mouse and human studies. Neural Regen Res. 2023; 18(3):521–522.

14. Klistorner A, Wang C, Yiannikas C, et al. Evidence of progressive tissue loss in the core of chronic MS lesions: A longitudinal DTI study. Neuroimage Clin. 2018; 17:1028–1035.

15. Klistorner S, Barnett MH, Klistorner A. Mechanisms of central brain atrophy in multiple sclerosis. MSJ. 2022; 28:2038–2045.

16. Rovira A, Auger C, Alonso J. Magnetic resonance monitoring of lesion evolution in multiple sclerosis. Ther Adv Neurol Disord. 2013; 6(5):298–310.

17. Coupé P, Mansencal B, Clément M, et al. AssemblyNet: A large ensemble of CNNs for 3D whole brain MRI segmentation. Neuroimage. 2020; 219:117–126.

18. Klistorner S, Barnett MH, Graham SL, Wang C, Klistorner A. Choroid plexus volume in multiple sclerosis predicts expansion of chronic lesions and brain atrophy. Ann Clin Transl Neurol. 2022; ahead of p.

19. Rodríguez-Lorenzo S, Konings J, Van Der Pol S, et al. Inflammation of the choroid plexus in progressive multiple sclerosis: Accumulation of granulocytes and T cells. Acta Neuropathol Commun. 2020; 8(1):1–13.

20. Rodríguez-Lorenzo S, Ferreira Francisco DM, Vos R, et al. Altered secretory and neuroprotective function of the choroid plexus in progressive multiple sclerosis. Acta Neuropathol Commun. 2020; 8(1):1–13.

21. Dixon GA, Pérez CA. Multiple Sclerosis and the Choroid Plexus: Emerging Concepts of Disease Immunopathophysiology. Pediatr Neurol. 2020; 103:65–75.

22. Vercellino M, Votta B, Condello C, et al. Involvement of the choroid plexus in multiple sclerosis autoimmune inflammation: A neuropathological study. J Neuroimmunol. 2008; 199(1–2):133–141.

23. Tonietto M, Poirion E, Lazzarotto A, et al. Periventricular remyelination failure in multiple sclerosis: a substrate for neurodegeneration. Brain. 2023; 146(1):182–194. Available at: https://doi.org/10.1093/brain/awac334.

24. Ricigliano VAGA, Morena E, Colombi A, et al. Choroid Plexus Enlargement in Inflammatory Multiple sclerosis. Radiology. 2021; 301(1):166–177. Available at: https://pubmed.ncbi.nlm.nih.gov/34254858/.

25. Barkho BZ, Monuki ES. Proliferation of cultured mouse choroid plexus epithelial cells. PLoS One. 2015; 10(3):1–14.

26. Campbell G, Kraytsberg E, Krishnan K, et al. Clonal Expansion of Mitochondrial DNA Deletions in Multiple Sclerosis. Acta Neuropathol. 2012; 124(1):209–220. Available at: https://www.ncbi.nlm.nih.gov/pmc/articles/PMC3624763/pdf/nihms412728.pdf.

27. Cardia E, Molina D, Abbate F, et al. Morphological modifications of the choroid plexus in a rodent model of acute ventriculitis induced by gram-negative liquoral sepsis – Possible implications in the pathophysiology of hypersecretory hydrocephalus. Child’s Nervous System. 1995; 11(9):511–516.

28. Wang X, Zhu Q, Yan Z, et al. Enlarged choroid plexus related to iron rim lesions and deep gray matter atrophy in relapsing-remitting multiple sclerosis. Mult Scler Relat Disord. 2023; 75(March):104740. Available at: https://doi.org/10.1016/j.msard.2023.104740.

29. Poirion E, Tonietto M, Lejeune FX, et al. Structural and Clinical Correlates of a Periventricular Gradient of Neuroinflammation in Multiple Sclerosis. Neurology. 2021; 96(14):e1865–e1875.

30. Magliozzi R, Howell OW, Reeves C, et al. A Gradient of neuronal loss and meningeal inflammation in multiple sclerosis. Ann Neurol. 2010; 68(4):477–493.

31. Magliozzi R, Scalfari A, Pisani AI, et al. The CSF Profile Linked to Cortical Damage Predicts Multiple Sclerosis Activity. Ann Neurol. 2020; 88(3):562–573.

32. Kuhlmann T, Moccia M, Coetzee T, et al. Multiple sclerosis progression: time for a new mechanism-driven framework. Lancet Neurol. 2022; 4422(22). Available at: http://www.ncbi.nlm.nih.gov/pubmed/36410373.

33. Klistorner SA, Barnett MH, Graham SL, Wang C, Klistorner A. The expansion and severity of chronic MS lesions follows a periventricular gradient. MSJ. 2022; (ahead of print).

34. Pardini M, Brown JWL, Magliozzi R, Reynolds R, Chard DT. Surface-in pathology in multiple sclerosis: a new view on pathogenesis? Brain. 2021.

35. Mainero C, Louapre C, Govindarajan ST, et al. A gradient in cortical pathology in multiple sclerosis by in vivo quantitative 7 T imaging. Brain. 2015; 138(4):932–945.

36. Magliozzi R, Fadda G, Brown RA, et al. “ Ependymal-in “ Gradient of Thalamic Damage in Progressive Multiple Sclerosis. Ann Neurol. 2022; 92:670–685.

37. Fadda G, Brown RA, Magliozzi R, et al. A surface-in gradient of thalamic damage evolves in pediatric multiple sclerosis. Ann Neurol. 2019; 85(3):340–351.

