## Supplementary material for "Longitudinal enlargement of choroid plexus is associated with inflammation and neurodegeneration in RRMS patients"

1. **Specific MRI parameters and image processing.**

The following MRI sequences were acquired:

a. Pre- and post-contrast (gadolinium) Sagittal 3D T1: GE BRAVO sequence, duration 4 min each, FOV 256mm, Slice thickness 1mm, TE 2.7ms, TR 7.2ms, Flip angle 12°, Pixel spacing 1mm. Acquisition Matrix (Freq x Phase) is 256x256, which results in 1mm isotropic acquisition voxel size. The reconstruction matrix is 256x256.
b. FLAIR CUBE; GE CUBE T2 FLAIR sequence, duration 6 min, FOV 240mm, Slice thickness 1.2 mm, Acquisition Matrix (Freq x Phase) 256x244, TE 163ms, TR 8000ms, Flip angle 90°, Pixel spacing 0.47 mm. The reconstruction matrix is 512x512.
c. Echo-Planar Imaging based diffusion weighted MRI, duration 9 min (64-directions with 2 mm isotropic acquisition matrix, TR/TE = 8325/86 ms, b = 1000 s/mm^2^, number of b0s = 2).

**MRI image pre-processing:**

The baseline T1-weighted imaging was realigned to Anterior and Posterior Commissure (AC-PC) orientation. Using FLIRT (FSL, FMRIB Software Library), follow-up T1 images were co-registered to initial (month 0) AC-PC space by applying transformation matrices derived from linear co-registration between baseline AC-PC aligned brain and follow-up native T1 brain images. In parallel, diffusion MRI was corrected for motion and eddy-current distortion in FSL, then EPI susceptibility distortion was minimized by applying deformation maps generated from nonlinear co-registration between DWI b0 brain images and T1-weighted images at each time-point using ANTS (Advanced Normalization Tools). Subsequently, tensor reconstruction was performed in MRtrix3. Tensor and FLAIR images were then linearly co-registered to corresponding T1 AC-PC images at each timepoint.

**DTI data processing:**

Diffusion weighted MRI data (dMRI) were pre-processed using tools provided by the software suites MRtrix3, FSL, and ANTs. Specifically, dMRI data were first denoised, then potential Gibbs-ringing artefacts were removed. The dMRI data were then corrected for bias field inhomogeneities using the ANTs N4 algorithm. a dMRI brain mask was then estimated using BET and used as an input, alongside the dMRI data, to eddy in order to correct for subject movement in the acquisition. Finally, phase distortion correction was applied using a non-linear registration method outlined below.

To correct for phase distortion within the dMRI data, first the brain was segmented from the corresponding T1w dataset, and a single B0 volume was extracted from the dMRI dataset. The T1w brain was then used as a mask to invert the contrast of the T1w image. A rigid-body registration was then performed on the inverted T1w image with the B0 volume as a target, which aligned the two images spatially. Non-linear registration was then performed using ANTs. The registration steps were comprised of a rigid body, then affine, and then SyN registration algorithm. The transformations and warps calculated from the non-linear registration steps were then applied to the entire dMRI dataset to correct for phase distortion artefacts.
